# Results and Impact of Intensive SARS-CoV-2 Testing in a High Volume, Outpatient Radiation Oncology Clinic in a Pandemic Hotspot

**DOI:** 10.1101/2020.08.11.20172551

**Authors:** Sean M McBride, Kimberly Bundick, Harper Hubbeling, Morgan Freret, Leslie A Modlin, Mini Kamboj, Oren Cahlon, Daniel R Gomez

**Affiliations:** Department of Radiation Oncology, Memorial Sloan Kettering Cancer Center, New York, USA; Department of Medicine, Division of Infectious Disease, Memorial Sloan Kettering Cancer Center, New York, USA

## Abstract

**Background:** In an attempt to reduce interruptions in radiation treatment, our department implemented universal SARS-CoV-2 PCR testing during the peak of the New York City COVID-19 epidemic.

**Methods:** Starting 4/18/20, outpatients coming into the Department of Radiation Oncology for either simulation or brachytherapy were required to undergo PCR testing for SARS-CoV-2. Starting on 5/6/20, patients were offered simultaneous SARS CoV-2 IgG antibody testing.

**Results:** Between 4/18/20-6/25/20, 1360 patients underwent 1,401 outpatient screening visits (Table 1). Of the patients screened, 411 were screened between 4/18/20 and 5/6/20 (*Phase 1*) with PCR testing: 13 (3.1%) patients were PCR positive. From 5/7/20 to 6/25/20, 990 patients were scheduled for both PCR and antibody testing (*Phase 2*), including 41 previously screened in Phase 1. Of those with known antibody status (n=952), 5.5% were seropositive. After 5/21/20, no screened patient (n=605) tested PCR positive. In the month prior to screening (3/17/20-4/19/20), 24 of 625 patients initiating external radiation had treatment interrupted due to COVID-19 infection (3.8%) vs 7 of 600 patients (1.1%) in the month post screening (4/20/20-5/24/20) (p=0.002).

**Conclusions:** State-wide mitigation efforts, coupled with intensive departmental screening, helped prevent interruptions in radiation during the COVID-19 epidemic that could have compromised treatment efficacy.

## Introduction

As of early August, there have been approximately 420,000 confirmed cases of COVID-19 with over 32,000 deaths in New York State (NYS). Owing to effective non-pharmaceutical interventions (NPI), daily case numbers in NYS declined from a peak of 10,000-11,000 cases/day in mid-April to 700-800 cases/day in late July.^1^ New York City began its phased re-opening on June 8^th^, 2020.^1^

Early in the pandemic, in order to limit radiation treatment interruptions and identify individuals in pre symptomatic stage of infection, we began a program of universal SARS-CoV-2 PCR testing and eventual antibody testing for all outpatients undergoing radiation simulation or brachytherapy. Herein we report a single institution case series with results from a broad-based screening program at a high-volume cancer center that maintained operations throughout the pandemic.

## Methods

Starting 4/18/20, outpatients coming into the Department of Radiation Oncology for either simulation or brachytherapy were required to undergo PCR testing for SARS-CoV-2. Starting on 5/6/20, patients were offered simultaneous antibody testing.

Patients underwent testing typically 24-72 hours before their simulation or brachytherapy. SARS-CoV-2 PCR testing was performed as previously described.^2^ Antibody testing was conducted using the Abbott SARS-CoV-2 IgG immunoassay.^3^ Patients were also screened for COVID-19 symptoms or contact with an infected individual, both the day prior and upon arrival for simulation or brachytherapy.

In addition to the test results, patient-specific variables (age, state of residence, insurance type, cancer type, symptom status at screening) were also collected; electronic chart review was used to determine whether patients with positive test results had, at any point since March 2020, reported symptoms consistent with COVID-19; radiation treatment interruptions were tracked with differences evaluated using the Chi-square test.

The database was closed on 6/25/20. This study was approved by the MSKCC Institutional Review Board (#20-216).

## Results

Between 4/18/20-6/25/20, 1360 patients underwent 1,401 outpatient screening visits (Table 1). Of the patients screened, 411 were screened between 4/18/20 and 5/6/20 (*Phase 1*) with PCR testing: 13 (3.1%) patients were PCR positive. From 5/7/20 to 6/25/20, 990 patients were scheduled for both PCR and antibody testing (*Phase 2*), including 41 previously screened in Phase 1. The results from this second screened population are presented in Table 2. After 5/21/20, no screened patient (n=605) tested PCR positive.

**Table 1:** Demographic and Disease Data.

|  | <u>Median</u> | <u>Range</u> |
| --- | --- | --- |
| Age (years) | 62 | 6-96 |
| Residence (State) | <u>No. (%)</u> | <u>%</u> |
| NY | 912 | 67.1 |
| NJ | 411 | 30.2 |
| CT | 18 | 1.3 |
| PA | 9 | 0.7 |
| Other | 10 | 0.7 |
| Cancer Diagnosis |  |  |
| Metastases | 411 | 30.2 |
| Breast | 403 | 29.6 |
| Genitourinary | 204 | 15.0 |
| Head and Neck | 80 | 5.9 |
| Gastrointestinal | 73 | 5.4 |
| Lung | 71 | 5.2 |
| Gynecologic | 47 | 3.5 |
| Lymphoma | 23 | 1.7 |
| CNS | 23 | 1.7 |
| Skin | 16 | 1.2 |
| Sarcoma | 9 | 0.7 |
| Insurance |  |  |
| Private | 771 | 56.7 |
| Medicare | 509 | 37.4 |
| Medicaid | 70 | 5.1 |
| Tricare | 5 | 0.4 |
| Uninsured | 3 | 0.2 |
| Self Pay | 2 | 0.1 |
| Gender |  |  |
| Male | 743 | 54.6 |
| Female | 617 | 45.4 |
|  | <u>n</u> | <u>%</u> |
| Symptomatic at Screening (Visits) |  |  |
| No | 1398 | 99.8 |
| Yes | 3 | 0.2 |

**Table 2:** Results from Phase 2 of Screening.

| **<u>Known (n=919)</u>** |  |
| --- | --- |
|  | **No. (%)** |
| **Ab Positive/PCR Positive** | **4 (0.4)** |
| **Ab Positive/PCR Negative** | **40 (4.3)** |
| **Ab Negative/PCR Positive** | **0** |
| **Ab Negative/PCR Negative** | **875 (95.2)** |

| **<u>Missing (n=71)</u>** |  |
| --- | --- |
|  | **No. (%)** |
| **Ab Positive/PCR Missing** | **8 (11.3)** |
| **Ab Negative/PCR Missing** | **25 (35.2)** |
| **Ab Missing/PCR Positive** | **0 (0)** |
| **Ab Missing/PCR Negative** | **38 (53.5)** |
| **% PCR Positive (of PCR known)** | **0.4%** |
| **% Ab Positive (of Ab known)** | **5.5%** |

abbreviations Ab=antibody, PCR=Polymerase Chain Reaction; percentage of patients who are PCR and antibody positive is calculated using a denominator that includes patients whose results were known (excludes missing).

In the month prior to screening (3/17/20-4/19/20), 24 of 625 patients initiating external radiation had treatment interrupted due to COVID infection (3.8%) vs 7 of 600 patients (1.1%) in the month post screening (4/20/20-5/24/20) (p=0.002).

Of the 17 patients with PCR positive screening tests, 7 (41.1%) had current or prior symptoms consistent with COVID-19; 16 completed antibody testing of which 12 (75.0%) were positive; for the 4 patients who were PCR +/ Ig G -, antibody testing occurred 13-42 days post positive PCR test.

Of the 52 patients seropositive at screening, 19 (36.5%) had current or prior symptoms consistent with COVID-19; 46 of the seropositive patients had simultaneous (within 2 days) PCR testing of which 4 (8.7%) were positive; 21 patients had PCR testing done > 2 days prior to antibody testing of which 10 (47.6%) were positive; in sum, all 52 seropositive patients had PCR testing done either at or prior to antibody screening of which 14 (26.9%) were positive. Of note, a symptomatic, PCR positive patient (PCR on 4/14/20), seropositive on 5/8/20, was found to be seronegative on repeat testing on 7/8/20.

## Discussion

Our data suggest that NPI initiated in late March contributed to a near elimination of COVID-19 in a vulnerable cancer population by late May. These mitigation efforts, coupled with our intensive screening, helped prevent interruptions in radiation that could have compromised treatment efficacy. Despite patients residing in areas of prior high endemicity, the seroprevalence in this cancer population was low suggesting persistent risk of infection and the need for continued vigilance.

## Data Availability

All relevant data is kept in a single, password protected database on an internal server.

